# Relative deprivation in income, tobacco chewing and bidi smoking among Indian adults: Findings from a cross-sectional national study

**DOI:** 10.1101/2021.06.12.21258833

**Authors:** Anupam Joya Sharma, Malavika Ambale Subramanyam

## Abstract

The robust relationship of income with health outcomes is widely documented and there is a rich body of work examining the direct impact of income on health. However, researchers have also extensively studied the relationship of an individual’s relative socioeconomic position with their wellbeing, frequently explained by the relative deprivation (RD) hypothesis. Despite the high socioeconomic inequality, research on RD in the Indian context is scarce. Therefore, the primary objective of this study was to assess the relationship of RD in income with the risk of tobacco chewing and bidi smoking in a sample of Indian adults. We drew data from the second round of the nationally representative India Human Development Survey for our analysis. Using data on annual household equivalized income from 26,529 adults, we created the Yitzhaki index to operationalize RD in income. We then fitted survey adjusted logistic regression models accounting for age, gender, absolute household equivalized income, education, place of residence, caste, and religion. Odds ratios from fully adjusted models for the third tertile (greatest relative deprivation) versus the first tertile (lowest relative deprivation) ranged between 1.32 (95% CI=1.15, 1.50) and 1.63 (95% CI=1.40, 1.89) for tobacco chewing and varied between 0.99 (95% CI=0.86, 1.14) and 1.32 (95% CI=1.15, 1.51) for bidi smoking, depending on the reference group used to compute the RD measure. Our findings suggested that with higher RD, the risk of consumption of smokeless tobacco and bidi smoking was higher. These results point to the health hazard of relative deprivation in a society that has enjoyed rapid economic growth coupled with rising income inequality in the last few decades. Our findings call for a comprehensive assessment of the socioeconomic inequality in income and health observed in India and the implementation of efficient programs to narrow the gap.

**Data availability statement:** The data that support the findings of this study are available in IHDS 2 at https://ihds.umd.edu/data/ihds-2.

## Introduction

The robust relationship of income with health outcomes is widely documented in the US (1) as well as India (2). With greater income, individuals have a greater access to health-preserving resources (3). Further, wealthier individuals also benefit from their higher social status, whereas the poor often suffer from stress due to their low social status and comparison with their “upper” class counterparts (4). Evidence suggests that the health protection offered by a higher socioeconomic status is observed throughout the socioeconomic spectrum and not restricted to a poor versus non-poor dichotomy (5). Therefore, beyond the direct impact of income on health, researchers have also extensively studied the relationship of an individual’s relative socioeconomic position with their wellbeing (6). One theory explaining this phenomenon is the theory of Relative Deprivation propounded by Runciman (7). According to this theory, individuals make upward social comparisons, consequently, the relatively deprived individuals (with less income relative to some reference group) may suffer from stress, frustration, and perceived injustice. The logic of relative deprivation lies in the idea that all human beings have an innate tendency to make upward social comparisons. Researchers have argued and showed evidence that such upward comparisons are linked to poorer health. A comprehensive review showcases the previous studies that have provided evidence for the relative deprivation hypothesis (8). Moreover, there is evidence that greater RD is related to adverse mental health outcomes (9,10), poor self-rated health (11–13), and risky-health behaviors such as smoking (13–15). Furthermore, in one study, premature mortality was also associated with higher levels of RD in income, independent of absolute income in Sweden (16).

Runciman’s (7) relative deprivation theory has been frequently cited to explain this complex relationship of RD with health status. For instance, RD likely limits individuals’ access to resources and services such as employment, as well as their social networks, thereby influencing their health-preserving opportunities. Furthermore, increased relative inequality could produce stress in the forms of shame or frustration among those who perceive being relatively deprived, possibly leading to poorer health outcomes.

While RD and its health-impacts have been extensively studied in high-income countries such as USA and UK, research on RD is scarce in middle- and low-income countries, especially ones with high income inequality such as India. Exploring relative income deprivation in India could be worthwhile for several reasons. Firstly, despite India being one of the fastest growing economies in the world, it has a wide income inequality (17,18). Due to such widened income inequalities, individuals sharing similar sociodemographic characteristics could be positioned differently on the wide income spectrum. Such inequalities within a similar group of individuals might lead to social comparisons among them, which according to the RD theory could increase the risk of poor health. Secondly, the implementation of programs in India offering avenues for upward social mobility in turn makes the individual experience of relative deprivation more prominent in such contexts. India has a long history of social stratification, especially in the context of gender, caste, and income. To tackle such inequalities, the government of India introduced several social welfare policies, such as the Mahatma Gandhi National Rural Employment Guarantee Act (MNREGA), National Rural Livelihood Mission (NRLM) and so on for the socially disadvantaged. Despite such avenues for upward social mobility if an individual perceives that their social position is disadvantaged compared to similar others and that they deserve to achieve a higher socioeconomic position, RD could be heightened in such contexts. While these factors make India an apt site for understanding RD and its psychosocial impact, we could not locate any Indian empirical study examining the relationship of RD with behavioral and health outcomes. Given the high prevalence of adverse behavioral outcomes (such as alcohol consumption and tobacco chewing)(19) and the inverse relationship between socioeconomic status and such outcomes in the Indian context, examining these outcomes through the RD pathway could have several policy implications in mitigating such outcomes. Therefore, the primary objective of this study was to assess the relationship of RD in income with the risk of tobacco chewing and bidi smoking in a sample of Indian adults. A previous study has shown a strong socioeconomic gradient in the consumption of smokeless tobacco in India (20). Relative deprivation, in addition to the absolute income, may explain the socioeconomic gradient in the tobacco chewing and bidi smoking in India. We hypothesized that greater levels of RD could lead to greater levels of frustrations and stress, thereby increasing the frequency of maladaptive behaviors such as tobacco chewing and bidi smoking among Indian adults.

Using the nationally representative 2012 India Human Development Survey (IHDS)-II data (21), we examined the association of relative deprivation in income with chewing tobacco and bidi smoking among Indian adults. We hypothesized that greater levels RD would be related to higher risk of tobacco chewing and bidi smoking even after accounting for the absolute socioeconomic measures such as income and education.

## Methods

### Data source and the sample

We drew data from the second round of the IHDS (2011-12) (21), one of the few nationally representative demographic surveys in India. The dataset is focused on the employment, health, family welfare, and social issues in the Indian context. Data for the second round were collected from 42,152 households in 1503 villages and 971 urban neighborhoods across India. A multi-stage random stratified sampling was employed. We removed observations with missing information regarding tobacco chewing and smoking bidi from our analyses. Data for all the required variables was available for only 26,529 individuals which comprised our analytical sample.

### Outcome variable

We chose two outcomes in our analysis. One of the outcome variables was a binary indicator of whether an individual had reported consumption of smokeless tobacco products (i.e., tobacco chewing) during the second wave of the IHDS survey. The second variable was a binary indicator of whether or not an individual had reported smoking bidi. Responses to these in the survey were recorded as “never,”, “rarely,” “sometimes,” and “daily.” For our analyses, we created consumption of smokeless tobacco and bidi smoking variables by collapsing the responses “rarely,” “sometimes,” and “daily” to “1” and the option “never” to “0.”

### Predictor variables

Relative deprivation (RD): We calculated the widely cited Yitzhaki index (22) to measure relative deprivation in income. This index is calculated based on the accumulated deficit between one’s own income and the incomes of all others in a social comparison group. We hypothesized that annual household income through salaries and wages (with bonuses) would yield a comprehensive income measure apt for measuring RD. Therefore, we used the “household income through salaries and wages” variable from the IHDS-2 dataset to create the RD variable. The average annual household income earned through salaries and wages in our sample was approximately 80000 Indian Rupees (INR) (range: 100 - 926000 INR). We equivalized this by following the Luxembourg Income study approach and divided the annual income with the square root of the total number of household members (23). We further converted it into units of 10000 INR (∼$135). Following Yitzhaki (22), we operationalized RD in income for each participant *i* as the average of the income difference between participant *i* and the remaining participants reporting a greater income than the participant *i* in their reference group. This is given by the formula— Relative deprivation (RD)= 1/N ∑ (y_j_ – y_i_), ∀ y_j_ □>□ y_i_, where individual *j* has a greater income than *i*.

Since conceptualizing reference group is often challenging, especially in a country such as India with diverse social identities associated with individuals, we drew on concepts from sociological theories on social comparison of individuals with “similar others” (24) to design our reference groups. Previous studies from the US had formed the reference groups based on combinations of different sociodemographic factors such as age, gender, education, and so on (12,15,25). Following this approach, we created 32 reference groups, with the combinations of the sociodemographic variables age (18-30 years, 31-49 years, 50-60 years, 61-99 years), gender (male, female), education (no education, up to 5^th^ grade, completed 5^th^ grade but not 10^th^ grade, completed 10^th^ grade but not graduated college, graduated college and above), caste (general category, reserved category), and religion (Hindu, Muslim, others). This included considering the whole sample as one reference group. The RD variables were then converted to RD tertiles.

We also created another predictor variable defining the position of an individual in the household income hierarchy within a reference group and operationalized it as quintiles. We estimated the influence of these ranks on the two outcomes separately for the reference groups which had the weakest and the strongest estimates of the association of RD with tobacco chewing.

### Covariates

We included age (in years), gender (male, female), place of residence (rural, urban), education (no education, up to 5^th^ grade, completed 5^th^ grade but not 10^th^ grade, completed 10^th^ grade but not graduated college, graduated college and above), caste (general category (privileged castes), reserved category (underprivileged castes)), and religion (Hindu, Muslim, others) as covariates in our analyses. We also accounted for the equivalized absolute annual household income from salaries and wages (in units of 10000 INR), because our initial attempts of using tertiles of the absolute annual household income variable resulted in a lack of statistical power.

### Statistical analyses

First, descriptive statistics were computed to assess the prevalence of tobacco chewing and bidi smoking across different social groups. The distribution of RD variables was then summarized across the social categories. Next, we employed survey adjusted logistic regression models to assess the relationship of RD (in tertiles) with tobacco chewing and bidi smoking (binary). Separate models were run for each of the 32 reference groups for each of the two outcomes. First, we adjusted the models for only age, gender, and household equivalized income (Model 1). Next, the logistic regression models were further adjusted for education, place of residence, caste, and religion (Model 2). Additionally, we fitted fully adjusted logistic regression models with the income rank quintile variable as the predictor for both the outcomes. For each outcome, two models were fit: one for the reference group which showed the weakest association of RD with chewing and the other which showed the strongest association.

Since the survey was conducted using a multistage stratified sampling procedure, all the models accounted for the complex sampling design of the data. *Alpha* was set at 0.05. Adjusted odds ratios are presented along with their 95% confidence intervals. All the models were run in Stata version 12.

### Ethical considerations

This study uses publicly available community-based dataset (21). It was a collaborative project of the University of Maryland, College Park, the National Council of Applied Economic Research (NCAER) in Delhi, Indiana University, and the University of Michigan. Informed consent was taken from all the respondents of this survey and interviews were conducted following all ethical protocols. Since our study is a secondary data analysis of publicly available de-identified data, this is considered exempt from ethics approval by the Institutional Ethics Committee of our university. More details about the IHDS can be found elsewhere (https://ihds.umd.edu/about).

## Results

The sociodemographic characteristics of the sample and the prevalence of tobacco chewing and bidi smoking across these characteristics are presented in Table 1. The mean age of the respondents in our sample was 45.5 years and mostly comprised men (∼83%). Notably, almost 75% of the sample comprised rural residents. While the prevalence of tobacco chewing was higher among women (∼82%) compared to men (∼66%), the prevalence of smoking bidi was higher among men (∼40%), versus women (∼16%).

**Table 1:** Sociodemographic distribution of the study participants in our analytical sample (N=26,529)

| Sociodemographic factor | Relative frequency |
| --- | --- |
| <u>Gender</u> |  |
| Men | 83.0 |
| Women | 17.0 |
| <u>Education</u> |  |
| No education | 50.5 |
| Up to 5th grade | 20.4 |
| Completed 5<sup>th</sup> grade but not 10<sup>th</sup> grade | 30.9 |
| Completed 10<sup>th</sup> grade but not graduated college | 7.0 |
| Graduated college and above | 1.1 |
| <u>Caste</u> |  |
| General | 81.1 |
| Reserved | 18.9 |
| <u>Religion</u> |  |
| Hindu | 83.0 |
| Muslim | 11.4 |
| Others | 5.6 |
| <u>Place of residence</u> |  |
| Urban | 24.9 |
| Rural | 75.1 |

Our survey adjusted logistic regression analyses showed a positive association of RD in income with tobacco chewing that was statistically significant for all the reference groups (including the overall sample group). This statistically significant association was observed in models which were adjusted for age, gender, and household equivalized income as well as the fully adjusted models. The fully adjusted odds ratios for the third tertile (greatest relative deprivation) versus the first tertile (lowest relative deprivation) ranged between 1.32 (95% CI=1.15, 1.50) and 1.63 (95% CI=1.40, 1.89) depending on the reference group. There was also a gradient in the odds of tobacco chewing across the tertiles of RD, which suggested that with higher RD, the risk of consumption of smokeless tobacco was higher. We could not detect any pattern in the variation of the odds ratios (the relationship of RD with tobacco chewing) across reference groups. Table 2 presents the estimates (adjusted odds ratios and 95% confidence intervals) of the association of RD with chewing tobacco in the sample.

**Table 2:** Association (adjusted odds ratios, 95% confidence interval (CI)) of RD in income with tobacco chewing in India.

| Reference group | Model 1 |  | Model 2 |  |
| --- | --- | --- | --- | --- |
|  | <u>RD Tertile 2</u> | <u>RD Tertile 3</u> | <u>RD Tertile 2</u> | <u>RD Tertile 3</u> |
| - |  |  |  |  |
| Age | 1.30 (1.15 , 1.46) | 1.43 (1.26, 1.64) | 1.28 (1.14, 1.44) | 1.38 (1.21, 1.58) |
| Gender | 1.32 (1.16, 1.49) | 1.68 (1.45, 1.95) | 1.31 (1.16, 1.48) | 1.63 (1.39, 1.89) |
| Education | 1.27 (1.13, 1.43) | 1.63 (1.43, 1.86) | 1.24 (1.10, 1.40) | 1.52 (1.31, 1.76) |
| Caste | 1.36 (1.20, 1.54) | 1.64 (1.41 , 1.90) | 1.34 (1.19, 1.52) | 1.61 (1.37, 1.88) |
| Religion | 1.29 (1.15, 1.46) | 1.67 (1.43 , 1.94) | 1.29 (1.14, 1.45) | 1.63 (1.39, 1.90) |
| Age*Gender | 1.25 (1.12, 1.40) | 1.37 (1.20, 1.56) | 1.23 (1.10, 1.38) | 1.32 (1.15, 1.50) |
| Age*Education | 1.28 (1.15, 1.43) | 1.54 (1.36, 1.74) | 1.24 (1.11, 1.39) | 1.42 (1.24, 1.62) |
| Age*Caste | 1.27 (1.12, 1.44) | 1.38 (1.20, 1.59) | 1.25 (1.10, 1.42) | 1.34 (1.16, 1.54) |
| Age*Religion | 1.24 (1.11, 1.39) | 1.41 (1.23, 1.61) | 1.22 (1.09, 1.37) | 1.36 (1.18, 1.56) |
| Gender*Education | 1.26 (1.12, 1.41) | 1.64 (1.44, 1.87) | 1.23 (1.09, 1.38) | 1.52 (1.32, 1.76) |
| Gender*Caste | 1.34 (1.19, 1.51) | 1.63 (1.41, 1.89) | 1.33 (1.18, 1.50) | 1.60 (1.37, 1.86) |
| Gender*Religion | 1.28 (1.14, 1.44) | 1.65 (1.41, 1.94) | 1.27 (1.13, 1.43) | 1.61 (1.38, 1.89) |
| Education*Caste | 1.31 (1.17, 1.48) | 1.66 (1.45, 1.89) | 1.27 (1.13, 1.43) | 1.54 (1.34, 1.79) |
| Education*Religion | 1.29 (1.15, 1.45) | 1.62 (1.42, 1.84) | 1.26 (1.12, 1.42) | 1.51 (1.31, 1.74) |
| Caste*Religion | 1.32 (1.18, 1.48) | 1.61 (1.38, 1.88) | 1.31 (1.17, 1.47) | 1.61 (1.39, 1.87) |
| Age*Gender*Education | 1.30 (1.17, 1.46) | 1.56 (1.38, 1.78) | 1.27 (1.13, 1.42) | 1.45 (1.26, 1.66) |
| Age*Gender*Caste | 1.27 (1.14, 1.43) | 1.38 (1.21, 1.57) | 1.25 (1.12, 1.40) | 1.34 (1.17, 1.53) |
| Age*Gender*Religion | 1.25 (1.11, 1.39) | 1.39 (1.22, 1.59) | 1.22 (1.09, 1.37) | 1.34 (1.17, 1.53) |
| Age*Education*Caste | 1.28 (1.15, 1.44) | 1.53 (1.35, 1.74) | 1.23 (1.10, 1.38) | 1.40 (1.22, 1.61) |
| Age*Education*Religion | 1.26 (1.11, 1.41) | 1.53 (1.34, 1.73) | 1.22 (1.08, 1.37) | 1.42 (1.23, 1.63) |
| Age*Caste*Religion | 1.24 (1.10, 1.39) | 1.37 (1.19, 1.57) | 1.22 (1.08, 1.37) | 1.33 (1.15, 1.54) |
| Gender*Education*Caste | 1.32 (1.17, 1.49) | 1.68 (1.47, 1.91) | 1.28 (1.13, 1.44) | 1.56 (1.36, 1.80) |
| Gender*Education*Religion | 1.29 (1.14, 1.45) | 1.64 (1.43, 1.87) | 1.26 (1.12, 1.41) | 1.53 (1.33, 1.76) |
| Gender*Caste*Religion | 1.29 (1.14, 1.45) | 1.66 (1.42, 1.94) | 1.28 (1.14, 1.44) | 1.67 (1.43, 1.94) |
| Education*Caste*Religion | 1.29 (1.16, 1.45) | 1.62 (1.42, 1.85) | 1.26 (1.12, 1.41) | 1.53 (1.33, 1.76) |
| Age*Gender*Education*Caste | 1.28 (1.14, 1.44) | 1.55 (1.36, 1.76) | 1.23 (1.09, 1.39) | 1.42 (1.23, 1.64) |
| Age*Gender*Education*Religion | 1.25 (1.11, 1.41) | 1.55 (1.36, 1.77) | 1.21 (1.07, 1.36) | 1.44 (1.26, 1.66) |
| Age*Education*Caste*Religion | 1.26 (1.12, 1.42) | 1.51 (1.33, 1.71) | 1.21 (1.07, 1.37) | 1.40 (1.22, 1.60) |
| Age*Gender*Caste*Religion | 1.23 (1.10, 1.38) | 1.39 (1.22, 1.59) | 1.21 (1.09, 1.35) | 1.36 (1.19, 1.55) |
| Gender*Education*Caste*Religion | 1.28 (1.14, 1.14) | 1.64 (1.43, 1.88) | 1.24 (1.11, 1.39) | 1.56 (1.35, 1.79) |
| Age*Gender*Education*Caste*Religion | 1.25 (1.10, 1.41) | 1.51 (1.33, 1.72) | 1.20 (1.06, 1.36) | 1.40 (1.22, 1.60) |
| All (the entire sample) | 1.35 (1.19, 1.53) | 1.69 (1.46, 1.95) | 1.34 (1.18, 1.52) | 1.63 (1.40, 1.89) |

Legend: All the non-bolded figures are statistically significant at alpha 0.05. Bolded figures are not-statistically significant ( $p>0.05$ )

Model 1: Adjusted for equivalized annual household income, age, and gender.

Model 2: Adjusted for equivalized annual household income, age, gender, education, caste, religion, and place of residence

Supporting our hypothesis, the fully adjusted models showed that higher levels of RD in income was associated with greater odds of bidi smoking in almost all the reference groups (with two exceptions) (Table 3). However, in addition to weak estimates of the association for many of the reference groups, almost half of the results were not statistically significant.

**Table 3:** Association (adjusted odds ratios, 95% confidence interval (CI)) of RD in income with bidi smoking in India.

| Reference group | Model 1 |  | Model 2 |  |
| --- | --- | --- | --- | --- |
|  | RD Tertile 2 | RD Tertile 3 | RD Tertile 2 | RD Tertile 3 |
| Age | 1.34 (1.23, 1.55) | 1.47 (1.29, 1.69) | 1.22 (1.09, 1.37) | 1.32 (1.15, 1.51) |
| Gender | 1.22 (1.07, 1.38) | 1.21 (1.04, 1.41) | **1.09 (0.97, 1.23)** | **1.08 (0.93, 1.25)** |
| Education | **0.96 (0.85, 1.10)** | 0.74 (0.64, 0.86) | **1.07 (0.95, 1.20)** | **1.03 (0.90, 1.18)** |
| Caste | 1.23 (1.08, 1.41) | 1.17 (1.02, 1.36) | **1.11 (0.98, 1.26)** | **1.06 (0.92, 1.22)** |
| Religion | 1.17 (1.03, 1.34) | **1.01 (0.85, 1.20)** | 1.14 (1.01, 1.28) | **1.05 (0.91, 1.22)** |
| Age*Gender | 1.39 (1.23, .56) | 1.46 (1.27, 1.66) | 1.23 (1.10, 1.38) | 1.29 (1.13, 1.48) |
| Age*Education | **1.10 (0.98, 1.24)** | **0.97 (0.84, 1.12)** | 1.15 (1.02, 1.29) | 1.27 (1.10, 1.46) |
| Age*Caste | 1.38 (1.23, 1.55) | 1.44 (1.26, 1.65) | 1.23 (1.10, 1.37) | 1.29 (1.13, 1.48) |
| Age*Religion | 1.27 (1.22, 1.43) | 1.22 (1.05, 1.42) | 1.21 (1.08, 1.36) | 1.25 (1.10, 1.43) |
| Gender*Education | **0.96 (0.85, 1.10)** | 0.76 (0.66, 0.87) | **1.06 (0.94, 1.19)** | **1.04 (0.91, 1.19)** |
| Gender*Caste | 1.22 (1.07, 1.39) | 1.19 (1.02, 1.39) | 1.10 (0.97, 1.24) | 1.07 (0.92, 1.24) |
| Gender*Religion | 1.15 (1.01, 1.30) | **1.01 (0.85, 1.21)** | **1.11 (0.99, 1.25)** | **1.05 (0.90, 1.21)** |
| Education*Caste | **0.96 (0.84, 1.09)** | 0.73 (0.64, 0.85) | **1.05 (0.93, 1.19)** | **1.01 (0.88, 1.16)** |
| Education*Religion | **0.93 (0.82, 1.06)** | 0.70 (0.60, 0.81) | **1.06 (0.94, 1.19)** | **1.03 (0.90, 1.18)** |
| Caste*Religion | 1.22 (1.07, 1.39) | **0.93 (0.79, 1.10)** | **1.18 (1.04, 1.34)** | **0.99 (0.86, 1.14)** |
| Age*Gender*Education | **1.10 (0.97, 1.24)** | **0.95 (0.82, 1.09)** | 1.15 (1.01, 1.28) | 1.22 (1.06, 1.40) |
| Age*Gender*Caste | 1.38 (1.23, 1.55) | 1.44 (1.26, 1.66) | 1.23 (1.09, 1.37) | 1.29 (1.12, 1.47) |
| Age*Gender*Religion | 1.25 (1.11, 1.41) | 1.21 (1.05, 1.41) | 1.19 (1.06, 1.33) | 1.24 (1.08, 1.41) |
| Age*Education*Caste | **1.07 (0.95, 1.21)** | **0.93 (0.81, 1.07)** | **1.12 (0.99, 1.26)** | 1.22 (1.06, 1.40) |
| Age*Education*Religion | **1.04 (0.92, 1.18)** | **0.88 (0.77, 1.01)** | 1.13 (1.00, 1.27) | 1.23 (1.07, 1.41) |
| Age*Caste*Religion | 1.29 (1.15, 1.45) | 1.21 (1.05, 1.41) | 1.23 (1.10, 1.37) | 1.27 (1.11, 1.45) |
| Gender*Education*Caste | **0.93 (0.81, 1.06)** | 0.73 (0.63, 0.85) | **1.01 (0.89, 1.15)** | **0.99 (0.86, 1.14)** |
| Gender*Education*Religion | **0.95 (0.83, 1.08)** | 0.71 (0.61, 0.82) | **1.07 (0.95, 1.21)** | **1.03 (0.90, 1.18)** |
| Gender*Caste*Religion | 1.21 (1.07, 1.38) | **0.99 (0.83, 1.17)** | 1.17 (1.04, 1.32) | **1.03 (0.89, 1.20)** |
| Education*Caste*Religion | **0.97 (0.86, 1.10)** | 0.70 (0.60, 0.81) | **1.08 (0.96, 1.21)** | **1.02 (0.88, 1.17)** |
| Age*Gender*Education*Caste | **1.05 (0.93, 1.18)** | **0.90 (0.79, 1.04)** | **1.09 (0.97, 1.22)** | **1.16 (1.01, 1.34)** |
| Age*Gender*Education*Religion | **1.05 (0.93, 1.18)** | **0.88 (0.76, 1.01)** | **1.12 (1.00, 1.25)** | 1.19 (1.04, 1.37) |
| Age*Education*Caste*Religion | **1.03 (0.92, 1.16)** | **0.88 (0.76, 1.01)** | **1.11 (0.99, 1.24)** | 1.22 (1.06, 1.39) |
| Age*Gender*Caste*Religion | 1.27 (1.13, 1.43) | 1.19 (1.03, 1.39) | 1.21 (1.08, 1.35) | 1.23 (1.07, 1.40) |
| Gender*Education*Caste*Religion | **0.94 (0.83, 1.07)** | 0.70 (0.60, 0.81) | **1.04 (0.93, 1.17)** | 1.00 (0.87, 1.15) |
| Age*Gender*Education*Caste*Religion | **1.06 (0.94, 1.19)** | **0.90 (0.78, 1.03)** | 1.12 (1.00, 1.26) | 1.22 (1.07, 1.39) |
| All (the entire sample) | 1.23 (1.08, 1.41) | 1.20 (1.03, 1.40) | **1.10 (0.97, 1.25)** | **1.07 (0.93, 1.25)** |

Legend: All the non-bolded figures are statistically significant at alpha 0.05. Bolded figures are not-statistically significant ( $p > 0.05$ )

Model 1: Adjusted for equivalized annual household income, age, and gender.

Model 2: Adjusted for equivalized annual household income, age, gender, education, caste, religion, and place of residence

Further, our fully adjusted models also showed a negative association of quintile ranks in income with tobacco chewing for the two reference groups which had the weakest and the strongest estimates for RD (Figure 1). However, we could find such a pattern for smoking bidi only for one reference group.

**Figure 1:**
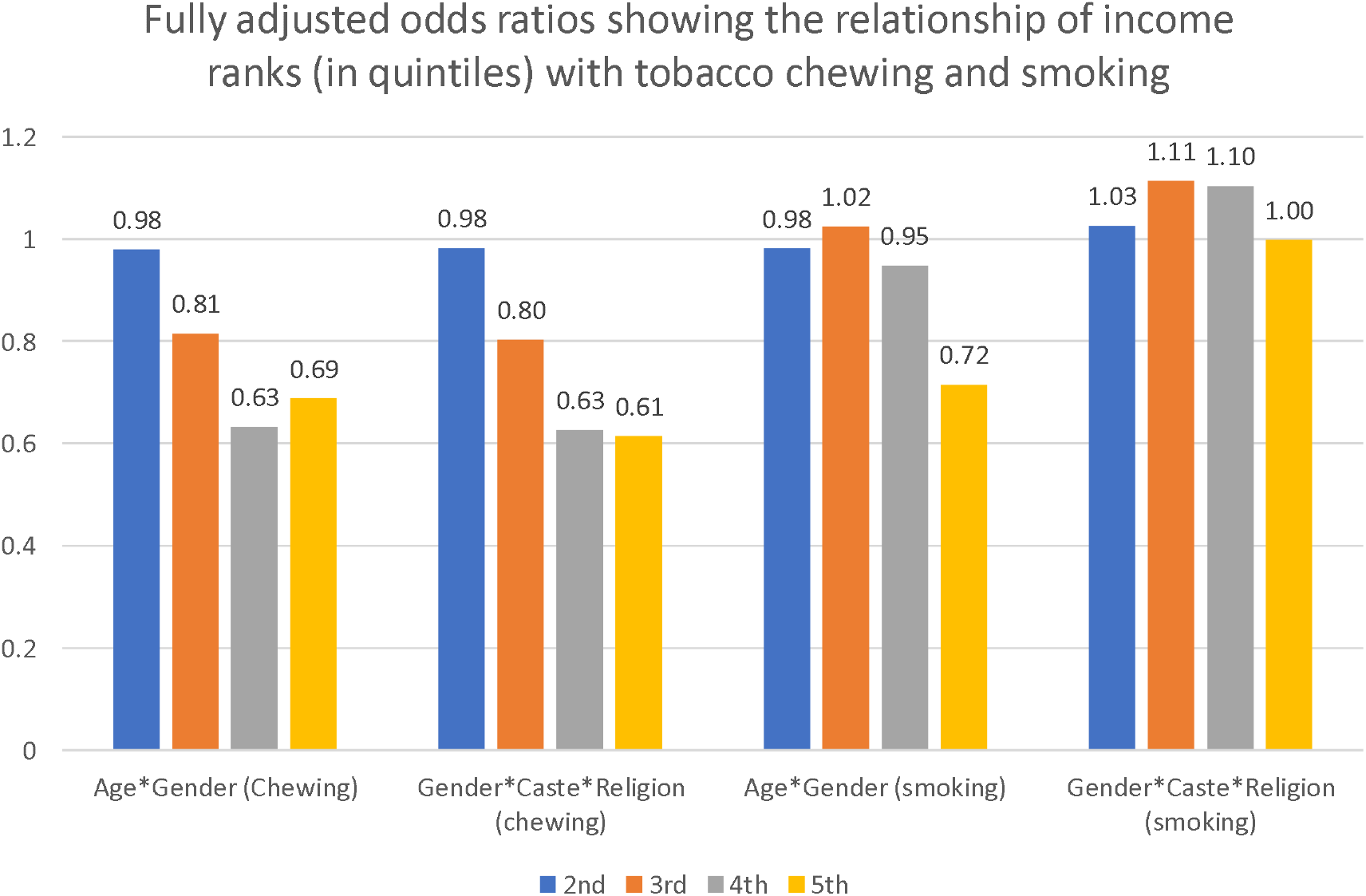
Bar graph showing the fully adjusted odds ratios for the relationship of income ranks (in quintiles) with tobacco chewing and smoking in two reference groups: Ange*Gender and Age*Caste*Religion

Our results from the adjusted models also showed that the absolute annual household income (equivalized) was negatively associated with tobacco chewing in all the models and the estimates were statistically significant. The adjusted odds ratios (for 10000 INR difference in income) ranged from 0.97 (95% CI=0.96, 0.99) to 0.99 (95% CI=0.97, 0.99). This suggests that with higher absolute income, the risk of chewing tobacco is lower among Indian adults, independent of their RD and sociodemographic factors. Our fully adjusted models also revealed that compared to men, women were at higher risk of tobacco chewing, independent of the sociodemographic factors and their relative deprivation.

## Discussion

Our findings from a nationally representative survey provide evidence for the relative deprivation hypothesis in India. We found that in individuals with low, versus high, relative deprivation in income, the prevalence of chewing tobacco was higher. This relationship was consistent even after accounting for the sociodemographic characteristics and equivalized absolute household income of the individuals. The estimates were almost similar across all the reference groups used in our study and also between both the models (with and without expanded list of covariates). However, we found limited evidence to support the hypothesized association for bidi smoking. While our final models suggested that the influence of RD in income on bidi smoking supported our hypothesis, a substantial proportion of the results lacked strong estimates and adequate statistical power. Notably, the finding that the ranking within a reference group in income, independent of the absolute income, was associated with tobacco chewing and bidi smoking further strengthens the support for the RD hypothesis. To our knowledge, this is the first study from India to assess the relationship of RD with the behavioral outcome tobacco chewing and bidi smoking, using a large community-based sample of Indian adults.

Our findings corroborate the findings from previous Western and Chinese studies which found that higher levels of RD were related to maladaptive health behaviors and outcomes. For instance, similar to our findings, one 2009 study from China (26) and another 2013 study from Taiwan (27) found that higher levels of RD was associated with higher health risk behavior such as increased smoking consumption. Further, research from the US has previously shown that relative deprivation was associated with mortality from tobacco-related cancers (a 58% increase) (15). While we could not assess the relationship of RD with mortality due to limited data, our finding of the relationship of higher levels of RD with greater odds of tobacco chewing supports the conclusions from the Eibner & Evans study (15). While previous studies of RD have focused on maladaptive behaviors such as smoking, we chose tobacco chewing and bidi smoking as our outcomes because they exhibited a clear socioeconomic gradient in the Indian population. However, there is no evidence of a strong socioeconomic gradient in cigarette smoking among Indian adults.

Previous research discusses one possible mechanism of health impacts of RD using the psychosocial pathway (28,29). For instance, the social comparison theory by Festinger (30) posits that, human beings make social comparisons and evaluate their position in the society, and in the absence of any benchmark they often compare themselves with similar others (reference groups), which could lead to a sense of deprivation. Thus, higher levels of RD in a reference group could lead to higher psychological stress among the deprived individuals thereby resulting in maladaptive behaviors such as smoking, drinking, and substance abuse (12). Another explanation relies on the access to material resources which may not be basic needs for sustenance but are required to live with dignity in a society (31). With the advancement of consumerism in India, individuals heavily rely on material resources (such as the Internet and mobile phones) which are beyond the basic needs of life. Such a consumerist change may create new standards of living in a society (32) and could deprive the individuals who could not afford such “luxurious” necessities from leading a fully engaged life, resulting in their poorer health outcomes and maladaptive health behaviors (12,16). For instance, the cheaper price of internet network plans in India have made internet affordable in India for a majority of the society, possibly creating new norms for improved standard of living. Therefore, such changes in standard of living in societies, for instance, increased use of mobile phones and the Internet, likely leads to the excessive reliance on such necessities in the dissemination of health-related information and provision of healthcare. For instance, mobile health initiatives by the Indian government in the dissemination of health-related information has increased over the last few years (33). Furthermore, such changes could also heighten the sense of deprivation among the marginalized who frequently do not have access to mobile phones or the Internet. This could enhance their stress likely increasing their risk of poorer health behaviors and outcomes.

While our findings for both tobacco chewing and bidi smoking were consistent in supporting our hypothesis, the strengths of the estimates and statistical power to measure them were not similar across both the outcomes. We found a statistically weaker association of RD in income with bidi smoking versus tobacco chewing in our sample. One explanation for such unexpected results could be the social and cultural norms around these two different types of tobacco consumption. For instance, in different parts of India, there is stigma associated with tobacco smoking (for example, bidi smoking). On the other hand, the practice of tobacco chewing has been viewed as culturally acceptable in India. In fact, tobacco is an integral part of several traditional items in India, such as *paan*, and has been perceived as part of the culture with minimal restriction in its consumption, even by the historically underprivileged groups such as women and the youth. We were unable to perform a deeper dive into the differing sociocultural determinants of tobacco chewing versus smoking given data restrictions.

### Limitations

The study has a few limitations that the readers need to bear in mind while interpreting the results. First, we did not have data on reference groups and therefore we cannot make claims about how an individual from India makes social comparisons. However, we have followed widely cited social theories on whom an individual makes a self-comparison with (30), for instance, individuals from similar social groups (same caste, age, neighborhood and so on). Therefore, our choice of reference groups following previous studies (12,16)-- all combinations of the sociodemographic factors age, gender, education, caste, and religion— makes our findings robust. In fact, we could not find any pattern in the change of estimates depending on the complexity of combinations of sociodemographic factors to define the reference groups. Second, we assumed that relatively deprived individuals are at higher risk of emotional stress, which might not be the case always. Protective inter-personal attributes such as positive personality and resilience could influence the levels of RD in a person. While material pathway of RD causing adverse behavior could explain our results in such cases, we argue that it is not always true since there could be a role of the emotional stress pathway as well. However, we do not have the data to provide evidence to distinguish the influence of both pathways separately. Third, while our sample was drawn from a national community-based survey, the sample comprised mostly men and rural residents. This limits the generalizability of our findings to all Indian adults. However, given sufficient diversity in our sample with respect to age, caste, and religion, we are confident that our results are informative and provide a comprehensive picture of RD, at least among the men from rural India. Lastly, the cross-sectional nature of our data limits our ability to establish causality. However, the new findings of RD and its relationship with tobacco chewing in the Indian context are informative. Despite these limitations, the strengths of the study are in its novel contribution to the Indian RD literature and its use of a large community-based sample of Indian adults. Further, the strong estimates of the relationship of RD with chewing of tobacco even after accounting for absolute household income makes our study findings robust.

### Implications and conclusion

Our findings call for a comprehensive assessment of the socioeconomic inequality in income and health observed in India and the implementation of efficient programs to narrow the gap. Further, these results point to the need for designing and delivery of behavior change programs that are sensitive to the potentially relatively deprived position of the intended program beneficiaries, beyond acknowledging their absolutely deprived position. Moreover, large scale quantitative studies are needed to examine the different pathways of how RD affects health. This would include the plan to collect data on potential mediators on these pathways. Qualitative studies are also warranted to explore the concept of social comparison in the Indian context, where individuals with diverse social identities cohabitate.

## Data Availability

The data that support the findings of this study are available in IHDS 2 at https://ihds.umd.edu/data/ihds-2.

## References

1. Subramanian S v., Belli P, Kawachi I. The macroeconomic determinants of health. Annual Review of Public Health. 2002.

2. Corsi DJ, Subramanian S v. Socioeconomic Gradients and Distribution of Diabetes, Hypertension, and Obesity in India. JAMA network open [Internet]. 2019 Apr 5 [cited 2020 Oct 20];2(4):e190411. Available from: /pmc/articles/PMC6450330/?report=abstract

3. Nobles J, Weintraub MR, Adler NE. Subjective socioeconomic status and health: Relationships reconsidered. Social Science and Medicine. 2013;82.

4. Wilkinson RG. Unhealthy Societies. Unhealthy Societies. 2002.

5. Ferrie JE, Shipley MJ, Davey Smith G, Stansfeld SA, Marmot MG. Change in health inequalities among British civil servants: The Whitehall II study. Journal of Epidemiology and Community Health. 2002;56(12).

6. Adjaye-Gbewonyo K, Kawachi I. Use of the Yitzhaki Index as a test of relative deprivation for health outcomes: A review of recent literature. Social Science and Medicine. 2012.

7. Runciman WG. Relative Deprivation and Social Justice. Journal of Personality & Social Psychology. 1966;

8. Adjaye-Gbewonyo K, Kawachi I. Use of the Yitzhaki Index as a test of relative deprivation for health outcomes: A review of recent literature. Vol. 75, Social Science and Medicine. 2012.

9. Wildman J. Income related inequalities in mental health in Great Britain: Analysing the causes of health inequality over time. Journal of Health Economics. 2003;

10. Kondo N, Saito M, Hikichi H, Aida J, Ojima T, Kondo K, et al. Relative deprivation in income and mortality by leading causes among older Japanese men and women: AGES cohort study. Journal of Epidemiology and Community Health. 2015;69(7).

11. Gravelle H, Sutton M. Income, relative income, and self-reported health in Britain 1979-2000. Health Economics. 2009;

12. Subramanyam M, Kawachi I, Berkman L, Subramanian S v. Relative deprivation in income and self-rated health in the United States. Social Science and Medicine. 2009;

13. Caner A, Yiğit YC. Relative deprivation and its association with health indicators: Lower inequality may not improve health. SSM - Population Health. 2019;7.

14. Ling DC. Do the Chinese “Keep up with the Jones”?: Implications of peer effects, growing economic disparities and relative deprivation on health outcomes among older adults in China. China Economic Review. 2009;

15. Eibner C, Evans WN. Relative deprivation, poor health habits, and mortality. Journal of Human Resources. 2005;

16. Yngwe MÅ, Lundberg O. Assessing the contribution of relative deprivation to income differences in health. In: Health Inequalities and Welfare Resources: Continuity and Change in Sweden. 2006.

17. India: extreme inequality in numbers | Oxfam International [Internet]. [cited 2020 Oct 20]. Available from: https://www.oxfam.org/en/india-extreme-inequality-numbers

18. Kurian NJ. Widening Regional Disparities in India_Some Indicators. Economic and Political Weekly. 2000;

19. IIPS. National Family Health Survey (NFHS-4) 2015-16, Factsheet, India. Ministry of Health and Family Welfare, Government of India. 2017.

20. Subramanian S v., Nandy S, Kelly M, Gordon D, Smith GD. Patterns and distribution of tobacco consumption in India: Cross sectional multilevel evidence from the 1998-9 national family health survey [Internet]. Vol. 328, British Medical Journal. BMJ Publishing Group; 2004 [cited 2020 Nov 15]. p. 801–6. Available from: http://www.bmj.com/

21. Vanneman R. India Human Development Survey. Human Development. 2008.

22. Yitzhaki S. Relative deprivation and the gini coefficient. Quarterly Journal of Economics. 1979;

23. Buhmann B, Rainwater L, Schmaus G, Smeeding TM. EQUIVALENCE SCALES. WELL□BEING, INEQUALITY, AND POVERTY: SENSITIVITY ESTIMATES ACROSS TEN COUNTRIES USING THE LUXEMBOURG INCOME STUDY (LIS) DATABASE. Review of Income and Wealth. 1988;

24. Singer E. Reference Groups and Social Evaluations. In: Social Psychology [Internet]. Routledge; 2019 [cited 2020 Nov 16]. p. 66–93. Available from: https://www.taylorfrancis.com/

25. Wu N, Fu A, Zhang Z, He W, Yao T, Sun X, et al. Relationship between relative deprivation and health of Hainan Island residents: Mediating effect of negative health behaviors. PeerJ. 2020;

26. Ling DC. Do the Chinese “Keep up with the Jones”?: Implications of peer effects, growing economic disparities and relative deprivation on health outcomes among older adults in China. China Economic Review. 2009;20(1).

27. Kuo CT, Chiang T liang. The association between relative deprivation and self-rated health, depressive symptoms, and smoking behavior in Taiwan. Social Science and Medicine. 2013;89.

28. Dawson DA, Grant BF, Ruan WJ. The association between stress and drinking: Modifying effects of gende and vulnerability. Alcohol and Alcoholism [Internet]. 2005 Sep 1 [cited 2020 Nov 22];40(5):453–60. Available from: https://academic.oup.com/alcalc/article/40/5/453/188522

29. Slopen N, Kontos EZ, Ryff CD, Ayanian JZ, Albert MA, Williams DR. Psychosocial stress and cigarette smoking persistence, cessation, and relapse over 9-10 years: A prospective study of middle-aged adults in the United States. Cancer Causes and Control. 2013;

30. Festinger L. A Theory of Social Comparison Processes. Human Relations. 1954;

31. Debes R. Adam smith on dignity and equality. Vol. 20, British Journal for the History of Philosophy. 2012.

32. Internet users in India: For the first time, India has more rural net users than urban | India Business News - Times of India [Internet]. [cited 2020 Nov 25]. Available from: https://timesofindia.indiatimes.com/business/india-business/for-the-first-time-india-has-more-rural-net-users-than-urban/articleshow/75566025.cms

33. m-Health | National Health Portal Of India [Internet]. [cited 2021 Jun 12]. Available from: https://www.nhp.gov.in/miscellaneous/m-health

